# A large-scale online survey of patients and the general public: Preferring safe and noninvasive neuromodulation for mental health

**DOI:** 10.1101/2024.01.10.24301043

**Authors:** Cyril Atkinson-Clement, Andrea Junor, Marcus Kaiser

## Abstract

While neurotechnology provides opportunities for novel mental health interventions, preferences of patients and the general public, and the reasons for their choices are still unclear. Here, we conducted a large-scale online survey with 784 participants, half of them suffering from psychiatric and/or neurological conditions. We asked about techniques ranging from invasive (pharmaceutical drugs and brain implants) to noninvasive approaches (ultrasound, magnetic, or electric stimulation). First, participants had a low level of prior knowledge but were interested and excited about these opportunities. Second, both patients and the general population preferred focused ultrasound stimulation (first choice) while drugs and implants were ranked 3^rd^ and 5^th^ out of five, respectively. Finally, that preference was mainly driven by whether they perceived a technology as safe, rather than as effective. Overall, this survey identifies safety as main criterion for interventions and shows a preference for novel noninvasive approaches such as focused ultrasound neuromodulation.

**Highlights:**

- Focused ultrasound is the preferred intervention for brain and mental health conditions
- The preference of techniques was consistent between healthy participants and patients suffering from brain or mental health conditions.
- Getting more information about different neuromodulation techniques reduced confusion and increased feelings of being excited, optimistic, and comfortable with these techniques
- The ranking of preference is linked with the perceived safety but less related with perceived efficiency of techniques indicating safety as more important criterion

## Introduction

Brain neuromodulation relates to the modification of brain functioning, mostly for a clinical aim to decrease diseases’ symptoms. Looking at the opinions of patients is crucial in assessing whether they will have a high degree of adherence to interventions or whether they will reject certain approaches straight away.

Due to advances in neurotechnology, a new generation of interventions for psychiatric and neurological conditions is becoming available ^1^. While patients and the public already know about pharmaceutical drugs and, to some extent, brain implants ^2^, others such as noninvasive electrical ^3^ or magnetic stimulation ^4^ might be affected by stereotypes (e.g. thinking about electro shocks). Yet other approaches, such as ultrasound stimulation ^5^, will be almost unknown. Such novel approaches for neuromodulation can be targeted to stimulate specific brain regions, unlike pharmaceutical drugs that affect the whole brain (and body). Moreover, noninvasive approaches remove the need for surgery compared to invasive interventions. Therefore, there is a potential to reduce negative side effects and/or invasiveness of interventions.

Population-based data can give an insight into what currently healthy participants, who could suffer from brain and mental health conditions in the future, think about different approaches. We are therefore not limited to participants that show up within the healthcare system. However, this makes such a population difficult to reach. We therefore chose the tool of an online survey to reach a population that is as large and diverse as possible. Surveys on different interventions often focus on treatment efficacy, e.g. in psychiatry ^6^ or on opinions of clinicians, e.g. concerning epilepsy surgery ^7^. While there are some surveys on patient opinions ^8^, they do not cover novel approaches such as noninvasive focused ultrasound neuromodulation. At the same time, ethical questions are relevant for both ultrasound ^9^ and other neuromodulation techniques ^10-12^.

Current surveys on opinions are limited to only including patients or clinicians, in focusing on a single brain or mental health condition, or in only evaluating one neuromodulation technology. For example, a meta-analysis on participants (n=163) undergoing transcranial magnetic stimulation (TMS) showed that TMS was perceived to be safe and beneficial, there were low levels of fear, and they were willing to recommend the intervention to others ^13^.

With our online survey, we aim to capture the representation of the general population (i.e., non-expert) about the general view of neuromodulation. We are particularly interested in the view on using neuromodulation for brain and mental health interventions, rather than for neuroenhancement in healthy persons ^14^. In this study, we look at opinions of healthy participants as well as patients concerning a range of interventions, from standard treatment (pharmaceutical drugs) and brain implants (e.g. deep-brain stimulation) to noninvasive approaches using electric, magnetic, or ultrasound stimulation.

## Results

### Participant characteristics

Overall, 784 participants submitted the online survey. About half of the participants were healthy (n=383, 48.8%) while about half suffered from psychiatric and/or neurological conditions (see Table 1). For all categories, there were more female than male participants (overall: 69.4% female).

**Table 1.** Sample characteristics.

| Categories | N | % |
| --- | --- | --- |
| Sex |  |  |
| Female | 544 | 69.4% |
| Male | 223 | 28.4% |
| Other / Prefer not to respond | 17 | 2.2% |
| Age |  |  |
| 18-29 | 162 | 20.7% |
| 30-44 | 199 | 25.4% |
| 45-59 | 184 | 23.5% |
| 60-74 | 185 | 23.6% |
| ≥75 | 54 | 6.9% |
| Country |  |  |
| UK | 734 | 93.6% |
| America | 18 | 2.3% |
| Asia | 14 | 1.8% |
| EU | 12 | 1.5% |
| Oceania | 6 | < 1% |
| Africa | 0 | 0% |
| Qualification level |  |  |
| High school diploma | 133 | 16.9% |
| Bachelor's degree | 287 | 36.6% |
| Master's degree | 164 | 20.9% |
| Doctoral degree | 77 | 9.8% |
| Other | 123 | 15.7% |
| Occupation |  |  |
| Unemployed, Students, pensioners, Casual workers | 311 | 39.7% |
| Middle management | 201 | 25.6% |
| Higher management | 127 | 16.2% |
| Office supervisors | 97 | 12.4% |
| Manual workers | 27 | 3.4% |
| Skilled manual workers | 21 | 2.7% |
| Annual income |  |  |
| Below £13,999 | 143 | 18.2% |
| £14,000 - £22,999 | 127 | 16.2% |
| £23,000 – £30,999 | 90 | 11.5% |
| £31,000 – £40,999 | 110 | 14% |
| £41,000 – £62,999 | 77 | 9.8% |
| Above £63,000 | 77 | 9.8% |
| Prefer not to respond | 160 | 20.4% |
| Brain condition |  |  |
| No | 383 | 48.8% |
| Neurological | 279 | 35.6% |
| Psychiatric | 64 | 8.2% |
| Neurological and Psychiatric | 58 | 7.4% |
| Knowledge about neuromodulation |  |  |
| 1-2 | 461 | 58.8% |
| 3-4 | 160 | 21.2% |
| 5-6 | 97 | 12.4% |
| 7-8 | 52 | 6.6% |
| 9-10 | 15 | 1.9% |

Most participants only had a limited prior knowledge of neuromodulation techniques (Figure 1A). On a scale from unfamiliar (1) to familiar (10), the average score was 2.75. The age of participants was distributed in a range from 18 to 86 years old (Figure 1B).

**Figure 1.**
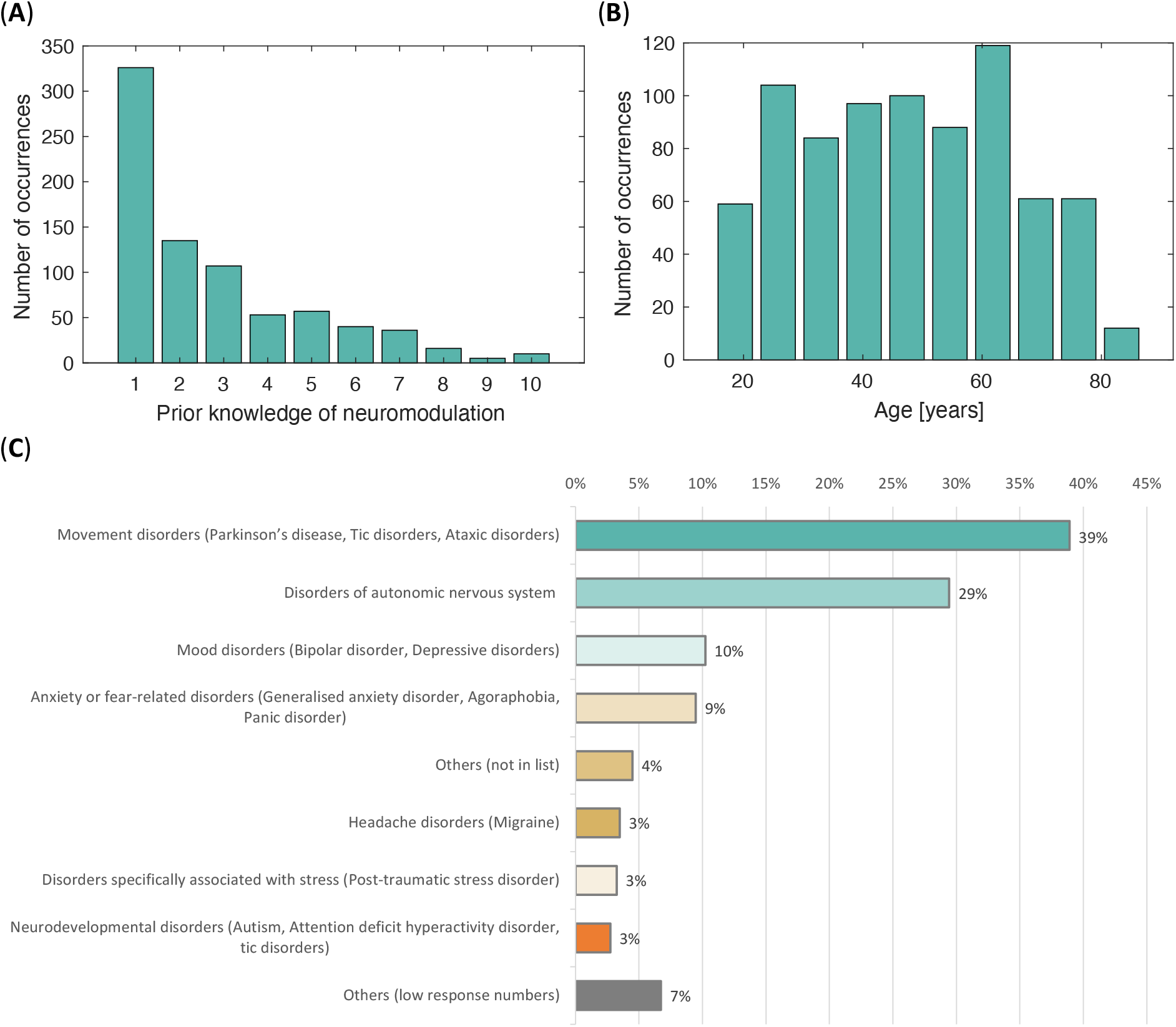
(**A**) Prior knowledge of neuromodulation ranging from 1 (lowest) to 10 (highest). Most participants had limited prior knowledge with an average score of 2.75. (**B**) Age of participants at the time of taking part in the survey. (**C**) Distribution of different psychiatric and neurological categories for different conditions. Others (low response number) summarizes conditions with six or fewer participants (1% or less of patients) while Others (not in list) indicates conditions that were not part of the options provided. Not that the total is above 100% as some participants indicated both neurological and psychiatric conditions.

We further tested for participants suffering from neurological and/or psychiatric conditions (Figure 1C). These participants indicated a range of conditions as their main condition with many reporting movement disorders (39%) and autonomous nervous system disorders (29%), but also conditions such as mood disorders (10%), anxiety (9%), and migraine (3%). Around 14% suffered from both neurological and psychiatric conditions.

In the following analysis, we only distinguish healthy participants, neurological condition only, psychiatric condition only, and both (neurological and psychiatric) conditions. We did not look at condition categories for two reasons. First, numbers are unequally distributed and too small for many condition categories. Second, condition categories contain several individual conditions, e.g. Parkinson’s, tics disorders, and ataxic disorders for the movement disorders category, which are again unequally distributed. However, separating along different conditions might be possible in the future if a larger sample size becomes available.

### Perceptions towards neuromodulation before and after receiving more information

Initially, people are mostly “interested” and “excited” about neuromodulation. After providing details about all different techniques, they are more “optimistic” (Chisq=175.7, p<0.001), “comfortable” (Chisq=69.2, p<0.001) and “excited” (Chisq=15.4, p<0.001), and less “interested” (Chisq=5.8, p=0.016) and “confused” (Chisq=63.4, p<0.001; Figure 2). No difference was found between healthy volunteers and patients (Chisq<2.6, p>0.45).

**Figure 2.**
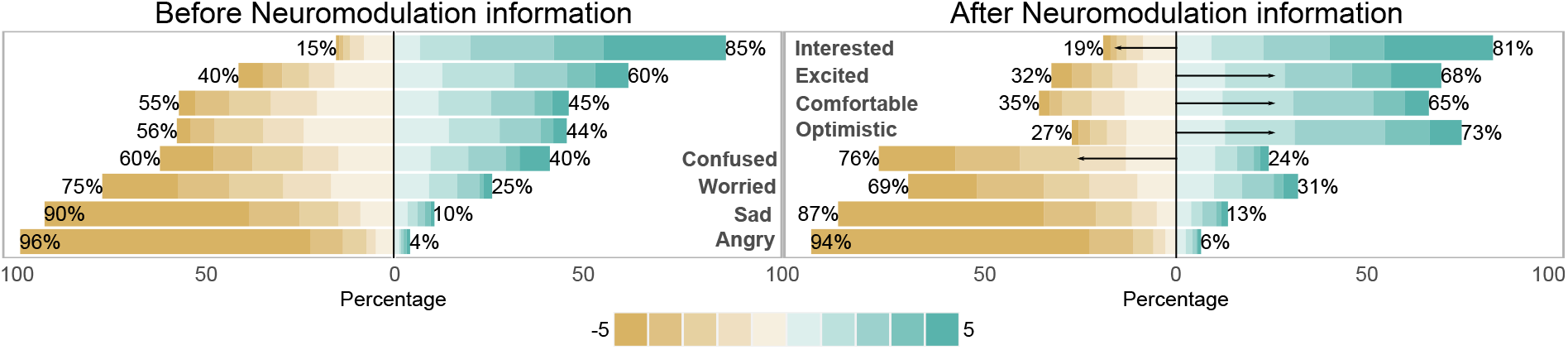
Perceptions of neuromodulation before (left) and after (right) receiving more information about different neuromodulation techniques. Significant changes after receiving information are indicated by arrows showing reduced (leftwards) and increased (rightwards) perceptions.

### Perception of different neuromodulation techniques

Neuromodulation approaches are differently preferred (Chisq=1015.1, p<0.001). All participants together, ultrasound was in the first position (score = 5.84) followed by magnetic stimulation and drugs (respectively 4.68 and 4.61, and not significantly different [p=0.97], followed by electrical stimulation (score = 4.16) and far below by brain implants (score = 3; Figure 3). However, we found some differences between healthy volunteers and patients (main effect, Chisq=30.6, p=0.002). In details, patients suffering from neurological conditions gave a higher preference score to drugs than healthy controls (p=0.0001), while ultrasound stimulation was preferred by psychiatric populations in comparison to the neurological one (p=0.021). All participants together, the degree of preference is highly due to the perceived safety of a technique (16.5% of the explained variance of the “preference”) and less by the perceived “effectivity” (6.2%).

**Figure 3.**
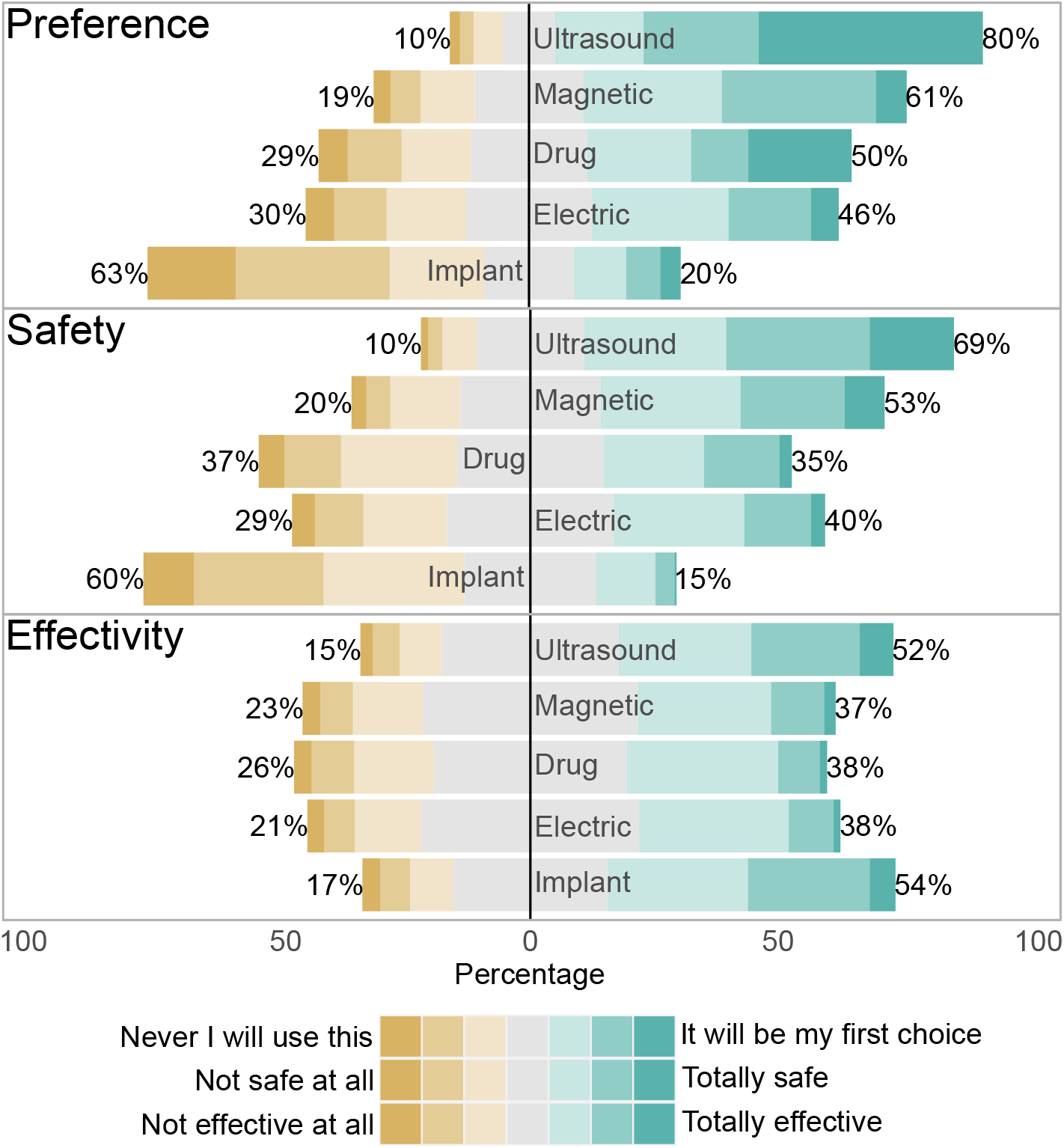
(top) Preference for different techniques ranging from ‘I will never use this’ to ‘It will be my first choice’, for different neuromodulation techniques: non-invasive ultrasound, magnetic, and electric stimulation, pharmaceutical drug interventions, and invasive brain implants. (middle) Perception of safety, ranging from ‘not safe at all’ to ‘totally safe’. (bottom) Perception of effectiveness (as in being able to improve brain and mental health conditions) for different techniques ranging from ‘Not effective at all’ to ‘Totally effective’.

### What does neuromodulation mean for you?

Giving participants the option to answer questions in their own words, after being given information about different techniques, we asked what neuromodulation means for them (“In just some words, what does neuromodulation means for you?”).

Looking at words indicating emotional valuation, we saw that most responses were positive – both for patients and healthy participants (Figure 4). Altogether, around 70% of answers were positive, 20% neutral, and 10% negative. Looking at different conditions, patients suffering from psychiatric conditions showed an even more positive evaluation than those suffering from neurological conditions. Within neurological conditions, Parkinson’s disease patients were also more positive, possibly being more aware of neuromodulation as a treatment option.

**Figure 4.**
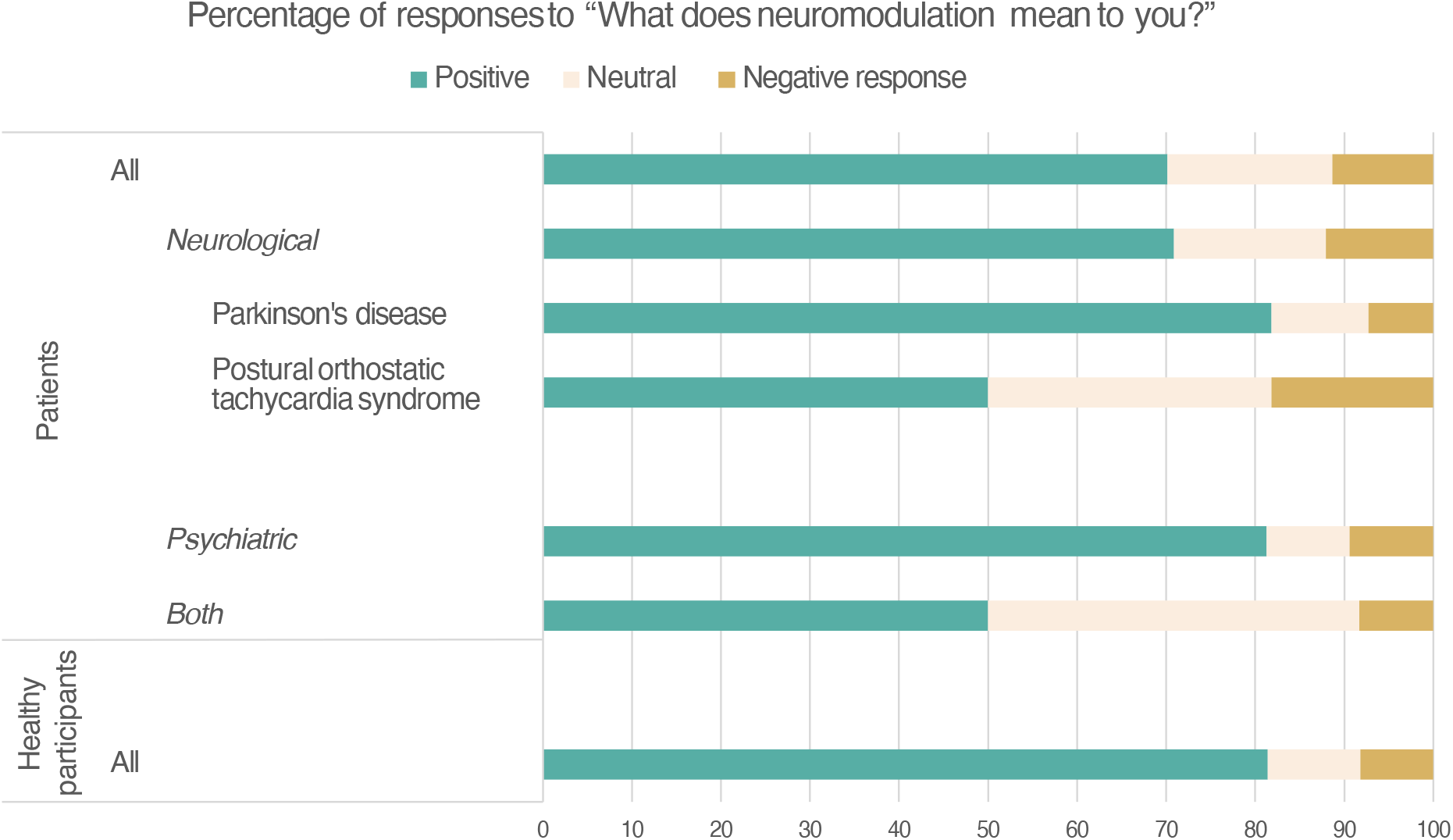
Emotional content of responses for the question “In just some words, what does neuromodulation means for you?”. Proportions of answers are given for patients and healthy participants. For patients, we also looked at sub-groups who suffered from neurological, psychiatric, or both conditions. Within the neurological category, we also looked at two specific conditions, Parkinson’s disease and Postural orthostatic tachycardia syndrome, for which a sufficient number of answers from patients were available.

Using data mining and cluster analysis of the corpus of answers, four topics were identified (Figure 5). First, neuromodulation for its clinical potential (words: “treatment”, “new”, “hope”) accounted for 64.4% of the corpus. Some characteristic answers here, for example, were: “Reminds me of Electroconvulsive therapy. Thought provoking. Maybe exciting with more research to help people”, “Possible future treatments but scary”, and “a new treatment for people who don’t have a choice”.

**Figure 5.**
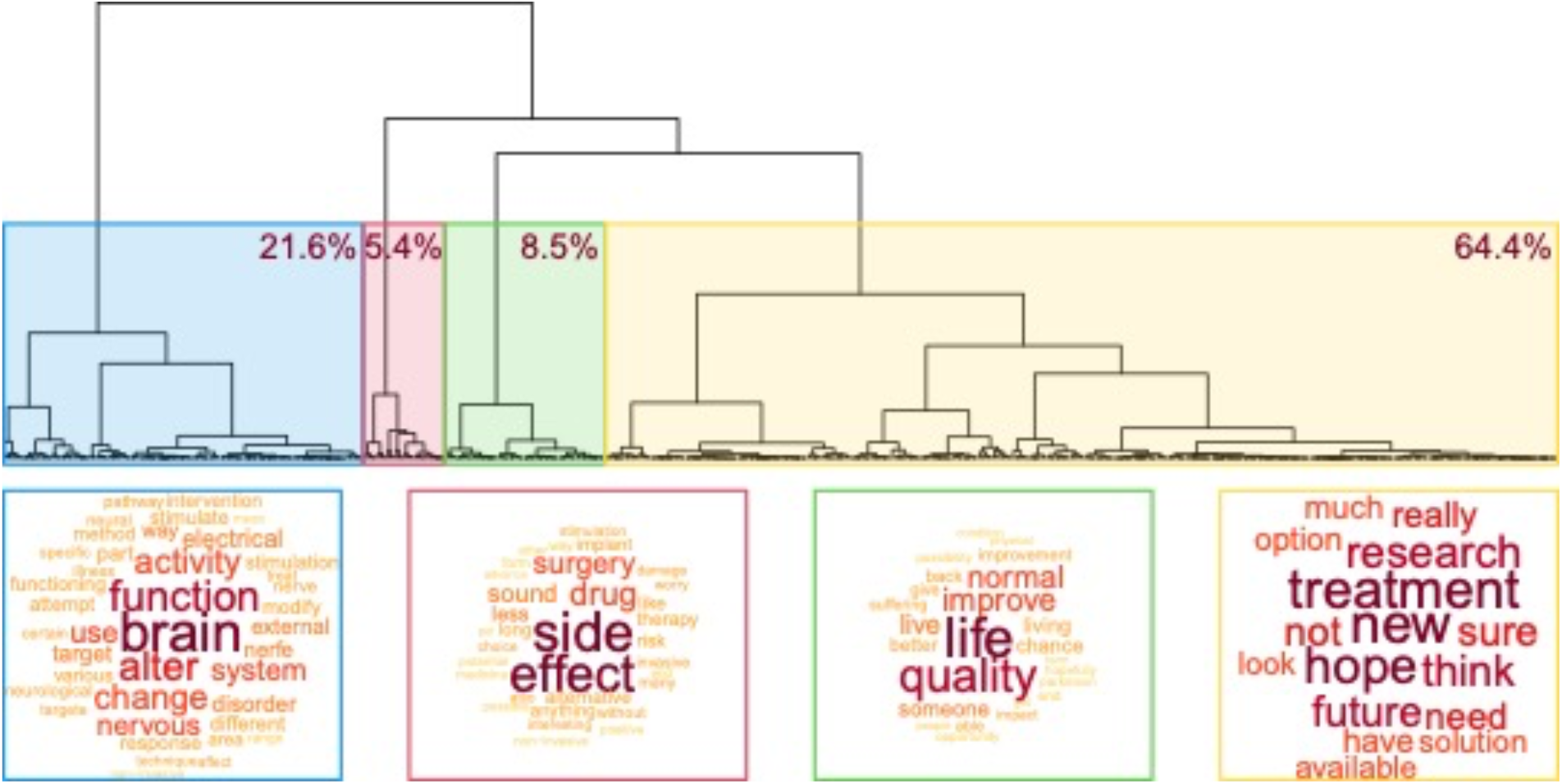
Topic analysis of the word corpus for the question “In just some words, what does neuromodulation means for you?”.

Second, neuromodulation as a definition (words: “function”, “brain”, “alter”, “change”) for 21.6% of the corpus, for example: “A way to alter brain activities”, “Neuromodulation means attempting to alter brain function in a favourable direction from the patient’s point of view”, “The alteration of brain electrical activity to support the individuals basic functioning”.

Third, neuromodulation for changing one’s life (words: “quality”, “life”, “improve”) for 8.5% of the corpus, for example: “Chance of living normal life or to maintain quality of life”, “Anything that can mask or improve my quality of life living with Parkinson’s disease must surely be explored”, and “A chance to give people a better standard of life. It’s not about how long we all live, but about the quality of life”.

Finally, potential concerns about neuromodulation (words: “side”, “effect”, “drug”) for 5.4% of the corpus, for example “Drugs are also not always very safe due to side effects so some alternatives like ultrasounds and electrical signals may be good”, “Drugs I worry about side effects, surgery I worry about any risks, and any of those signals transferred into my brain scare me too”, “Sounds like it has huge potential for non-invasive ways to treat conditions without drugs and avoid side effects”. Note that concerns were mainly raised for traditional interventions such as drug treatment.

## Discussion

We conducted a large-scale study with 784 participants to observe the opinions towards techniques that are already used to modulate neural activity (pharmaceutical drugs and, to a much lower extent, brain implants) as well as novel interventions including, for example, focused ultrasound stimulation. Around half of the participants suffered from psychiatric or neurological conditions. We find that participants had a low level of knowledge of neuromodulation techniques but were interested and excited about these opportunities. Getting more information about different neuromodulation techniques reduced confusion and increased feelings of being excited, optimistic, and comfortable with these techniques. Second, both patients and healthy participants preferred focused ultrasound stimulation (first choice), along with other noninvasive neuromodulation techniques such as transcranial magnetic stimulation. On the other hand, interventions such as drugs and implants were ranked 3^rd^ and 5^th^ out of five, respectively. Third, the preference for an intervention was mainly driven by whether they perceived a technology as safe, rather than whether they perceived it as effective. Finally, when asking about what neuromodulation means for them, answers indicated mostly (70%) positive feelings with only about 10% of negative responses – a result which showed no apparent differences between groups.

Our survey covers a comparatively high number of participants and is unique in looking at the perception of the general population as well, but results could be biased by the details we provide on each approach (see Supplemental Material for the provided details). Participants of this survey are geographically mostly limited to the United Kingdom. Moreover, the voluntary participants might not be representative of the general population or the population of patients suffering from brain and mental health issues with females over-represented in this survey ^15^. Finally, not all mental and brain health conditions are covered and, if covered, do not show the same prevalence as in the patient population. Nonetheless, given that results are similar between healthy participants and patients, one might expect that, at least for the conditions that are covered, perceptions of neuromodulation are comparable.

It would be interesting to have a larger representative study of the population in the UK and abroad. Moreover, we only looked at opinions of adults while the opinion of younger participants would be helpful in assessing noninvasive stimulation approaches for neurodevelopmental conditions. Finally, not all conditions are well represented, and charities might be interested in studying the opinion of patients in such cases.

Nonetheless, these results could have major implications for government and industry investment in mental health innovations. They indicate that safety rather than efficiency is a major concern. Consequently, novel interventions that are noninvasive, especially using ultrasound stimulation, will have fewer obstacles to face in terms of convincing patients and patients’ adherence to treatment. If demonstrated as effective for many conditions, such novel interventions could in the future change the way we are doing medicine, especially for neurological and psychiatric conditions. Therefore, non-invasive approaches should benefit of a higher attention (and funding) to reach the dual objective of both clinical usefulness and higher patients’ adherence.

## Conclusion

In conclusion, this study indicates that noninvasive approaches are considered safe and are preferred both by the general population and by people suffering from brain and mental health conditions. Safety is seen as more important than efficacy of interventions. Consequently, focused ultrasound neuromodulation that is seen as safest technique is also the preferred intervention. Overall, our insights into priorities and preferences of current and potential future patients could help in the decisions for future research directions in novel interventions for mental health.

## Methods

### Ethics

This online survey was approved by the University of Nottingham Faculty of Medicine & Health Sciences Research Ethics Committee (FMHS 147-1022). Participants confirmed that they were adults (>18 years) and agreed that they anonymised data would be used for this study following privacy and data handling standards (GDPR).

### Participant recruitment

Participants were recruited over a range of channels including Twitter/X, UK and international charities for brain and mental health conditions, and mailing lists. While most participants came from the United Kingdom, there were responses from around the world.

### Survey questionnaire

Completing the survey took around 10 minutes. The complete set of questions, including information given before agreeing to participate, are given in the supplementary information.

Participants were asked whether they suffered from one or more psychiatric conditions and could choose the most severe one from Neurodevelopmental disorders (Autism, Attention deficit hyperactivity disorder, tic disorders), Schizophrenia or other primary psychotic disorders (Delusional disorder, Schizoaffective disorder), Catatonia, Mood disorders (Bipolar disorder, Depressive disorders), Anxiety or fear-related disorders (Generalised anxiety disorder, Agoraphobia, Panic disorder), Obsessive-compulsive or related disorders (Hypocondriasis, Body dysmorphic disorder), Disorders specifically associated with stress (Post-traumatic stress disorder), Dissociative disorders, Feeding or eating disorders (Anorexia nervosa, Bulimia nervosa), Elimination disorders (Enuresis, Encopresis), Disorders of bodily distress or bodily experience, Disorders due to substance use or addictive behaviours, Impulse control disorders (Pyromania, Kleptomania, Intermittent explosive disorder), Disruptive behaviour or dissocial disorders, Personality disorders and related traits, Paraphilic disorders (Pedophilic disorder, Exhibitionistic disorder), Factitious disorders, Neurocognitive disorders (Delirium, Amnestic disorder, Dementia), Sleep-wake disorders (Insomnia, Hypersomnolence disorders), and Sexual dysfunctions.

They were also asked whether they (also) suffered from a neurological condition and could choose from Movement disorders (Parkinson’s disease, Tic disorders, Ataxic disorders), Disorders with neurocognitive impairment as a major feature (Alzheimer’s disease, Lewy body disease), Multiple sclerosis or other white matter disorders (Neuromyelitis optica, Leukodystrophies), Epilepsy or seizures, Headache disorders (Migraine), Cerebrovascular diseases (Stroke, Cerebral ischaemia), Spinal cord disorders excluding trauma (Myelopathy, Myelitis, Cauda equina syndrome), Motor neuron diseases or related disorders (Spinal muscular atrophy), Disorders of nerve root, plexus or peripheral nerves (Radiculopathy), Diseases of neuromuscular junction or muscle (Myasthenia gravis), Cerebral palsy, Nutritional or toxic disorders of the nervous system (caused by obesity or alcohol-use), Disorders of cerebrospinal fluid pressure or flow (Hydrocephalus), Disorders of autonomic nervous system, Human prion diseases (Creutzfeldt-Jacob disease), Disorders of consciousness, Postprocedural disorders of the nervous system (caused by surgery or medical process), Injuries of the nervous system, Neoplasms of the nervous system, Structural developmental anomalies of the nervous system, Syndromes with central nervous system anomalies as a major feature, Paralytic symptoms (Tetraplegia, Hemiplegia), and Dissociative neurological symptom disorder.

For both psychiatric and neurological conditions, participants had the option to select ‘I do not know’ or ‘Other’.

### Statistical analyses

All the statistical analyses were achieved using R. The threshold for significance was set at p≤0.05.

We used ordered logistic regressions (package MASS) in order to assess how providing information about neuromodulation change the degree of each agreement with the 8 adjectives we considered (interested, excited, comfortable, optimistic, confused, worried, sad, angry), and the degree of perceived preference, safety and effectivity of the 5 neuromodulation approaches we considered (ultrasound, magnetic and electrical stimulation, drugs and brain implants). To measure the role of the perceived safety and effectivity on the participants’ preference, we ran an ordered logistic regression which use both the safety and effectivity scores as independent variables and the preference score as dependent variable (after merging the responses for the 5 neuromodulation approaches). Then, we extracted the total variance and the part of explained variance for each of the predictor.

For the evaluation (positive/neutral/negative) of the responses to the question “what does neuromodulation mean to you?”, we separated answers into two groups according to whether or not the respondents identified themselves as having a neurological and/or psychiatric condition. Those who did not identify themselves as having a condition were categorised as “healthy participants”. Those who identified themselves as having a condition were further categorised according to the type of condition; neurological, psychiatric or both. For those within the neurological condition that identified as having either Parkinson’s disease or Postural orthostatic tachycardia syndrome we created two subgroups. Note that other conditions within the neurological condition existed, but their numbers were too small to include them as further sub-groups.

For further text analysis of the free text about the item “what neuromodulation means for you”, we first preprocessed the data as follow: i. each text was decomposed in smaller units called “segments” of approximately 20 words by respecting, when possible, sentences and punctuation; ii. each segment was lemmatized, which means that each word was transformed into its most basic form (no conjugation, no agreement); iii. words were selected based on the following criteria: they had to be composed of at least three letters, used by at least 1% of the participants, and be part of the following groups: adjectives, adpositions, adverbs, interjections, nouns, proper nouns, verbs (as tagged by the udpipe package). Punctuation, numbers, symbols or “stop words” (i.e., words of very high frequency and nonspecific meaning, e.g., “the”, “this”, “it” etc.) were not selected; iv. a binary matrix was built for each corpus (i.e., all segments extracted for one specific question), which included all text segments as rows and all selected words as columns. The number “1” was used when a word was used in a segment, while “0” was used when a word was missing in a segment.

Then, we applied Principal Component Analysis (PCA) to the raw binary matrices followed by Hierarchical Clustering on Principal Components (HCPC). The PCA was used to reduce the data complexity to only keep the most important information (i.e., the first five dimensions). The optimal number of clusters following HCPC was determined using the Ward’s criterion. This threshold is based on the selection of the option which leads to the highest relative loss of inertia (corresponding to the variance). The relation between all the clusters, words and segments were then measured as follows: for words, a *v.test* was applied to identify if a given word was more likely to be found in a specific cluster in comparison with the whole corpus. Each segment was assigned to one specific cluster. We also computed a specificity composite score based on the distance between the center of the cluster and the closest center of other clusters as follows:

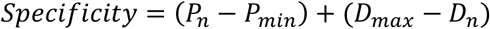

Where P corresponds to the proximity with the cluster center of the selected segment (*P*_*n*_), compared with the most central segment (*P*_*min*_), and D corresponds to the distance from the closest other cluster for the selected segment (*D*_*n*_) compared with the closest segment (*D*_*max*_). A low score means that the selected cluster is both close to the center of its cluster and far from the centers of other clusters.

## Data Availability

All data produced in the present study are available upon reasonable request to the authors

## Acknowledgements

M.K., C.A., and A.J. were supported by the Engineering and Physical Sciences Research Council (EP/W004488/1, EP/X01925X/1, and EP/W035057/1). M.K. was also supported by the Guangci Professorship Program of Rui Jin Hospital (Shanghai Jiao Tong University).

