## Supplementary Information for "A large-scale online survey of patients and the general public: Preferring safe and noninvasive neuromodulation for mental health"

### Neuromodulation survey

**Study Title:** Online survey to capture opinions about neuromodulation - **Research Team:** Pr. Marcus Kaiser, School of Medicine, University of Nottingham - **Faculty of Medicine & Health Sciences Research Ethics Ref: FMHS 147-1022**

This study is investigating how people perceive neuromodulation. You are invited to take part because you are aged >18 years. Please read through this information before agreeing to participate. You can ask any questions before deciding by contacting the researchers. Taking part is entirely voluntary.

**What will I be asked to do?**
After clicking the next button at the end of this information page you will be presented with a consent page. You will then be presented with some information and questions about neuromodulation. It should take you about 10 minutes to complete. No background knowledge is required. We would like you to answer all questions as honestly and completely as possible. You can withdraw at any point during the questionnaire for any reason, before submitting your answers by closing the browser. The data will only be uploaded by clicking the SUBMIT button on the final page.

**What are the disadvantages of taking part?**
It is possible that you may find some of the content in this questionnaire makes you feel uncomfortable. Please take time to think carefully about whether it might be a sensitive topic for you at the moment.

**What are the advantages of taking part?**
Your contribution will help the researchers to understand more about the different opinions regarding neuromodulation.

**Who will know I have taken part in the study?**
No one will know you have taken part in this study because we will not ask for your name or any other personal ID during this questionnaire, except if you contact us to ask further questions or choose to be contacted again about further studies. These will be received and handled separately from your completed questionnaire and it will not be possible to link the sets of data to protect your privacy. Your e-mail address will only be kept as long as needed to resolve your query and/or send you details of further studies. It will then be permanently deleted. For further information about how the university processes personal data please see: https://www.nottingham.ac.uk/utilities/privacy.aspx. Your IP address will not be visible to or stored by the research team. As with any online related activity the risk of breach is possible, but this risk is being minimized by using a platform that sits on an encrypted webpage.

**What will happen to your data?**
Your data will be stored in a folder sitting on a restricted access server at the University under the terms of its data protection policy. Data is kept for a minimum of 7 years. The results will be written up as an academic publications and scientific presentations. The overall anonymised data from this study may be shared for use in future research and teaching (with research ethics approval).

**Who will have access to your data?**
The University of Nottingham is the data controller (legally responsible for data security) and the Supervisor of this study is the data custodian (manages access to the data) and as such will determine how your data is used in the study. Your research and personal data will be used for the purposes of the research only. Research is a task that we perform in the public interest. Responsible members of the University of Nottingham may be given access to data for monitoring and/or audit of the study to ensure it is being carried out correctly. If you have any questions or concerns about this project, please contact: Dr. Cyril Atkinson-Clement or Dr. Marcus Kaiser. If you remain unhappy and wish to complain formally, you should then contact FMHS Research Ethics Commitee Administrator.

1. I have read and understood the above information and consent form, I confirm that I am 18 years old or older and by clicking the NEXT button to begin the online questionnaire, I indicate my willingness to voluntarily take part in the study. *

◯ I consent to take part

◯ I do not give consent

#### Thank you for participating!

Please, tick each box to continue:

##### 2. Question *

Please select 5 options.

- [ ] I confirm that I have read and understood the information on the previous page.
- [ ] I am 18 years old and/or older.
- [ ] I understand that my participation is voluntary, and I can end the study at any time and withdraw my data.
- [ ] I understand that my answers are anonymous.
- [ ] I understand the overall anonymized data from this study may be used in the future for research (with research ethics approval) and teaching purposes.

#### Some basic information about neuromodulation

*Neuromodulation is the alteration of nerve activity through targeted delivery of a stimulus to specific neurological sites in the body. It is carried out to change nervous tissue function.*

3. How much knowledge do you currently have about neuromodulation? *

*None at all* ☆ ☆ ☆ ☆ ☆ ☆ ☆ ☆ ☆ ☆ *Extensive*

4. Based only on the information provided above, how **optimistic** do you feel about neuromodulation? *

Not optimistic at all ☆ ☆ ☆ ☆ ☆ ☆ ☆ ☆ ☆ ☆ Very optimistic

5. Based only on the information provided above, how **comfortable** do you feel about neuromodulation? *

Not comfortable at all ☆ ☆ ☆ ☆ ☆ ☆ ☆ ☆ ☆ ☆ Very comfortable

6. Based only on the information provided above, how **sad** do you feel about neuromodulation? *

Not sad at all ☆ ☆ ☆ ☆ ☆ ☆ ☆ ☆ ☆ ☆ Very sad

7. Based only on the information provided above, how **excited** do you feel about neuromodulation? *

Not excited at all ☆ ☆ ☆ ☆ ☆ ☆ ☆ ☆ ☆ ☆ Very excited

8. Based only on the information provided above, how **worried** do you feel about neuromodulation? *

Not worried at all ☆ ☆ ☆ ☆ ☆ ☆ ☆ ☆ ☆ ☆ Very worried

9. Based only on the information provided above, how **confused** do you feel about neuromodulation? *

Not confused at all ☆ ☆ ☆ ☆ ☆ ☆ ☆ ☆ ☆ ☆ Very confused

10. Based only on the information provided above, how **angry** do you feel about neuromodulation? *

Not angry at all ☆ ☆ ☆ ☆ ☆ ☆ ☆ ☆ ☆ ☆ Very angry

11. Based only on the information provided above, how **interested** do you feel about neuromodulation? *

Not interested at all ☆ ☆ ☆ ☆ ☆ ☆ ☆ ☆ ☆ ☆ Very interested

#### More details and some scenarios involving neuromodulation

Now, we will describe five different technologies that induce change in brain function:
**1- Chemical method (drugs)** consists of ingesting a specific molecule to change brain function. This approach is often used as a first treatment for most brain diseases.

- Advantage: easy to consume.
- Disadvantages: invasive, does not target the specific brain area which is impaired, could induce numerous side effects, have generally to be consume during a long period or even for life, could induce an addiction.

**2- Brain implants** consists of implanting a device inside the brain to locally apply electrical stimulation and interrupt irregular brain signals that cause disease symptoms. This approach is used for diseases which are resistant to drugs or for which drugs induce significant side effects.

- Advantage: target the specific brain area which is impaired, could have a significant impact with few side effects, can be removed if strong side effects are experienced.
- Disadvantages: invasive, require neurosurgery which could be dangerous (risk of infection, neural immune system reactions, etc.), require a battery which must be recharged or replaced periodically, results are not immediate and require several months of parameterising.

**3- Electrical stimulation** uses direct electrical current to stimulate specific parts of the brain. A constant, low intensity current is passed through two electrodes placed over the head which modulates neuronal activity. This approach is currently under development.

- Advantage: non-invasive, painless.
- Disadvantages: it could induce transient side effects such as skin irritation, nausea, headache or dizziness, several treatment sessions repeated daily for several weeks are needed, does not target the specific brain area which is impaired.

**4- Magnetic stimulation** relies on electromagnetic induction using an insulated coil placed over the scalp, focused on an area of the brain. The coil generates brief magnetic pulses, which pass easily and painlessly through the skull and into the brain. The pulses generated are of the same type and strength as those generated by magnetic resonance imaging machines. This approach is currently under development.

- Advantage: non-invasive, painless.
- Disadvantages: it does not target the specific brain area which is impaired, several treatment sessions repeated daily for several weeks are needed.

**5- Ultrasound stimulation** consists of applying a low intensity ultrasonic energy with a transducer placed over the head through the brain to change neural activity. This approach is currently under development.

- Advantages: non-invasive, painless, target the specific brain area which is impaired.
- Disadvantages: several treatment sessions repeated daily for several weeks are needed.

12. In the future, if you have a condition which requires to choose between these approaches, what would be your preferences? (can you rank them from your first to your last choice?) *

- Chemical method (drugs)
- Electrical stimulation
- Ultrasound stimulation
- Magnetic stimulation
- Brain implants

13. In the future, if you have a condition which requires one of these approaches, what would be your opinion about the following approaches? *

| | 0 - Never I will use this | 1 | 2 | 3 | 4 | 5 | 6 - It will be my first choice |
|---|---|---|---|---|---|---|---|
| **Chemical method (drugs)** | ○ | ○ | ○ | ○ | ○ | ○ | ○ |
| **Brain implants** | ○ | ○ | ○ | ○ | ○ | ○ | ○ |
| **Electrical stimulation** | ○ | ○ | ○ | ○ | ○ | ○ | ○ |
| **Magnetic stimulation** | ○ | ○ | ○ | ○ | ○ | ○ | ○ |
| **Ultrasound stimulation** | ○ | ○ | ○ | ○ | ○ | ○ | ○ |

14. From your point of view, how **effective** do you think the following approaches are?
*

| | 0 - Not effective at all | 1 | 2 | 3 | 4 | 5 | 6 - Totally effective |
|---|---|---|---|---|---|---|---|
| **Chemical method (drugs)** | ○ | ○ | ○ | ○ | ○ | ○ | ○ |
| **Brain implants** | ○ | ○ | ○ | ○ | ○ | ○ | ○ |
| **Electrical stimulation** | ○ | ○ | ○ | ○ | ○ | ○ | ○ |
| **Magnetic stimulation** | ○ | ○ | ○ | ○ | ○ | ○ | ○ |
| **Ultrasound stimulation** | ○ | ○ | ○ | ○ | ○ | ○ | ○ |

15. From your point of view, how **safe** do you think the following approaches are?
*

| | 0 - Not safe at all | 1 | 2 | 3 | 4 | 5 | 6 - Totally safe |
|---|---|---|---|---|---|---|---|
| **Chemical method (drugs)** | ○ | ○ | ○ | ○ | ○ | ○ | ○ |
| **Brain implants** | ○ | ○ | ○ | ○ | ○ | ○ | ○ |
| **Electrical stimulation** | ○ | ○ | ○ | ○ | ○ | ○ | ○ |
| **Magnetic stimulation** | ○ | ○ | ○ | ○ | ○ | ○ | ○ |
| **Ultrasound stimulation** | ○ | ○ | ○ | ○ | ○ | ○ | ○ |

16. Based on the further details provided above, how **optimistic** do you feel about neuromodulation? *

Not optimistic at all ☆ ☆ ☆ ☆ ☆ ☆ ☆ ☆ ☆ ☆ Very optimistic

17. Based on the further details provided above, how **comfortable** do you feel about neuromodulation? *

Not comfortable at all ☆ ☆ ☆ ☆ ☆ ☆ ☆ ☆ ☆ ☆ Very comfortable

18. Based on the further details provided above, how **sad** do you feel about neuromodulation? *

Not sad at all ☆ ☆ ☆ ☆ ☆ ☆ ☆ ☆ ☆ ☆ Very sad

19. Based on the further details provided above, how **excited** do you feel about neuromodulation? *

Not excited at all ☆ ☆ ☆ ☆ ☆ ☆ ☆ ☆ ☆ ☆ Very excited

20. Based on the further details provided above, how **worried** do you feel about neuromodulation? *

Not worried at all ☆ ☆ ☆ ☆ ☆ ☆ ☆ ☆ ☆ ☆ Very worried

21. Based on the further details provided above, how **confused** do you feel about neuromodulation? *

Not confused at all ☆ ☆ ☆ ☆ ☆ ☆ ☆ ☆ ☆ ☆ Very confused

22. Based on the further details provided above, how **angry** do you feel about neuromodulation? *

Not angry at all ☆ ☆ ☆ ☆ ☆ ☆ ☆ ☆ ☆ ☆ Very angry

23. Based on the further details provided above, how **interested** do you feel about neuromodulation? *

Not interested at all ☆ ☆ ☆ ☆ ☆ ☆ ☆ ☆ ☆ ☆ Very interested

24. In just some words, what does neuromodulation means for you?

#### Some information about you

25. What is your sex? *

- ◯ Female
- ◯ Male
- ◯ Other
- ◯ I would prefer not to respond

26. What is your age (in years)? *

Number must be between 1 ~ 100

27. Which country do you live in? *

28. Which city do you live in? *

29. What is your highest level of qualification? *

- ◯ High school diploma
- ◯ Bachelor's degree
- ◯ Master's degree
- ◯ Doctoral degree
- ◯ Other

30. What is your occupation? *

- ◯ Higher management (e.g., bankers, lawyers, doctors)
- ◯ Middle management (e.g., teachers, creative and media people)
- ◯ Office supervisors (e.g., junior managers, nurses)
- ◯ Skilled manual workers (e.g., plumbers, builders)
- ◯ Semi-skilled and unskilled manual workers
- ◯ Unemployed, students, pensioners, casual workers

31. What is your annual income? *

- ◯ Below £13,999 (16,699€)
- ◯ Between £14,000 and £22,999 (16,700€ - 27,499€)
- ◯ Between £23,000 and £30,999 (27,500€ - 36,999€)
- ◯ Between £31,000 and £40,999 (37,000€ - 48,999€)
- ◯ Between £41,000 and £62,999 (49,000€ - 74,999€)
- ◯ Above £63,000 (75,000€)
- ◯ I would prefer not to respond

32. Do you take drugs for any conditions (except occasional use)? *

- ◯ Yes
- ◯ No

33. Have you ever had surgery (apart from basic surgery for wisdom tooth or appendicitis)? *

- ◯ Yes
- ◯ No

34. Do you suffer from a diagnosed psychiatric or neurological disease? *

- ◯ No
- ◯ Psychiatric
- ◯ Neurological
- ◯ Both

#### Some information about your psychiatric condition

35. Do you suffer from several diagnosed psychiatric conditions? *

- ○ Only one
- ○ Yes, more than one

36. What is the exact name of your condition? *

*If you suffer from several conditions, please consider only the one which is the most severe*

37. What is the classification of your condition? *

*If you suffer from several conditions, please consider only the one which is the most severe*

- ○ Neurodevelopmental disorders (Autism, Attention deficit hyperactivity disorder, tic disorders)
- ○ Schizophrenia or other primary psychotic disorders (Delusional disorder, Schizoaffective disorder)
- ○ Catatonia
- ○ Mood disorders (Bipolar disorder, Depressive disorders)
- ○ Anxiety or fear-related disorders (Generalised anxiety disorder, Agoraphobia, Panic disorder)
- ○ Obsessive-compulsive or related disorders (Hypocondriasis, Body dysmorphic disorder)
- ○ Disorders specifically associated with stress (Post-traumatic stress disorder)

- Dissociative disorders
- Feeding or eating disorders (Anorexia nervosa, Bulimia nervosa)
- Elimination disorders (Enuresis, Encopresis)
- Disorders of bodily distress or bodily experience
- Disorders due to substance use or addictive behaviours
- Impulse control disorders (Pyromania, Kleptomania, Intermittent explosive disorder)
- Disruptive behaviour or dissocial disorders
- Personality disorders and related traits
- Paraphilic disorders (Pedophilic disorder, Exhibitionistic disorder)
- Factitious disorders
- Neurocognitive disorders (Delirium, Amnestic disorder, Dementia)
- Sleep-wake disorders (Insomnia, Hypersomnolence disorders)
- Sexual dysfunctions
- I do not know
- Other

38. If you answered "Other" to the previous question, could you specify?

39. For how long have you suffered from this condition (from the diagnosis, in years)? *

*If you suffer from several conditions, please consider only the one which is the most severe*

The value must be a number

40. Do you receive treatment for this condition? *

*If you suffer from several conditions, please consider only the one which is the most severe*

- No, none
- Yes, drugs only
- Yes, surgery only
- Yes, brain stimulation (other than drugs)
- Yes, both drugs and surgery
- Yes, both drugs and brain stimulation (other than drugs)
- Yes, drugs, surgery and brain stimulation (other than drugs)
- Other

41. If you answered "Other" to the previous question, could you specify?

#### Some information about your neurological condition

42. Do you suffer from several diagnosed neurological conditions? *

- ◯ Only one
- ◯ Yes, more than one

43. What is the exact name of your condition? *

*If you suffer from several conditions, please consider only the one which is the most severe*

44. What is the classification of your condition? *

*If you suffer from several conditions, please consider only the one which is the most severe*

- ◯ Movement disorders (Parkinson’s disease, Tic disorders, Ataxic disorders)
- ◯ Disorders with neurocognitive impairment as a major feature (Alzheimer’s disease, Lewy body disease)
- ◯ Multiple sclerosis or other white matter disorders (Neuromyelitis optica, Leukodystrophies)
- ◯ Epilepsy or seizures
- ◯ Headache disorders (Migraine)
- ◯ Cerebrovascular diseases (Stroke, Cerebral ischaemia)
- ◯ Spinal cord disorders excluding trauma (Myelopathy, Myelitis, Cauda equina syndrome)

- Motor neuron diseases or related disorders (Spinal muscular atrophy)
- Disorders of nerve root, plexus or peripheral nerves (Radiculopathy)
- Diseases of neuromuscular junction or muscle (Myasthenia gravis)
- Cerebral palsy
- Nutritional or toxic disorders of the nervous system (caused by obesity or alcohol-use)
- Disorders of cerebrospinal fluid pressure or flow (Hydrocephalus)
- Disorders of autonomic nervous system
- Human prion diseases (Creutzfeldt-Jacob disease)
- Disorders of consciousness
- Postprocedural disorders of the nervous system (caused by surgery or medical process)
- Injuries of the nervous system
- Neoplasms of the nervous system
- Structural developmental anomalies of the nervous system
- Syndromes with central nervous system anomalies as a major feature
- Paralytic symptoms (Tetraplegia, Hemiplegia)
- Dissociative neurological symptom disorder
- I do not know
- Other

45. If you answered "Other" to the previous question, could you specify?

46. For how long have you suffered from this condition (from the diagnosis, in years)? *

*If you suffer from several conditions, please consider only the one which is the most severe*

The value must be a number

47. Do you receive treatment for this condition? *

*If you suffer from several conditions, please consider only the one which is the most severe*

- ◯ No, none
- ◯ Yes, drugs only
- ◯ Yes, surgery only
- ◯ Yes, brain stimulation (other than drugs)
- ◯ Yes, both drugs and surgery
- ◯ Yes, both drugs and brain stimulation (other than drugs)
- ◯ Yes, drugs, surgery and brain stimulation (other than drugs)
- ◯ Other

48. If you answered "Other" to the previous question, could you specify?

#### Some information about your psychiatric and neurological conditions

49. Do you suffer from several diagnosed psychiatric conditions? *

- ◯ Only one
- ◯ Yes, more than one

50. What is the exact name of your psychiatric condition? *

*If you suffer from several conditions, please consider only the one which is the most severe*

51. What is the classification of your psychiatric condition? *

*If you suffer from several conditions, please consider only the one which is the most severe*

- ◯ Neurodevelopmental disorders (Autism, Attention deficit hyperactivity disorder, tic disorders)
- ◯ Schizophrenia or other primary psychotic disorders (Delusional disorder, Schizoaffective disorder)
- ◯ Catatonia
- ◯ Mood disorders (Bipolar disorder, Depressive disorders)
- ◯ Anxiety or fear-related disorders (Generalised anxiety disorder, Agoraphobia, Panic disorder)
- ◯ Obsessive-compulsive or related disorders (Hypocondriasis, Body dysmorphic disorder)
- ◯ Disorders specifically associated with stress (Post-traumatic stress disorder)

- Dissociative disorders
- Feeding or eating disorders (Anorexia nervosa, Bulimia nervosa)
- Elimination disorders (Enuresis, Encopresis)
- Disorders of bodily distress or bodily experience
- Disorders due to substance use or addictive behaviours
- Impulse control disorders (Pyromania, Kleptomania, Intermittent explosive disorder)
- Disruptive behaviour or dissocial disorders
- Personality disorders and related traits
- Paraphilic disorders (Pedophilic disorder, Exhibitionistic disorder)
- Factitious disorders
- Neurocognitive disorders (Delirium, Amnestic disorder, Dementia)
- Sleep-wake disorders (Insomnia, Hypersomnolence disorders)
- Sexual dysfunctions
- I do not know
- Other

52. If you answered "Other" to the previous question, could you specify?

53. For how long have you suffered from this psychiatric condition (from the diagnosis, in years)? *

*If you suffer from several conditions, please consider only the one which is the most severe*

The value must be a number

54. Do you receive treatment for this psychiatric condition? *

*If you suffer from several conditions, please consider only the one which is the most severe*

- ◯ No, none
- ◯ Yes, drugs only
- ◯ Yes, surgery only
- ◯ Yes, brain stimulation (other than drugs)
- ◯ Yes, both drugs and surgery
- ◯ Yes, both drugs and brain stimulation (other than drugs)
- ◯ Yes, drugs, surgery and brain stimulation (other than drugs)
- ◯ Other

55. If you answered "Other" to the previous question, could you specify?

56. Do you suffer from several diagnosed neurological conditions? *

- ◯ Only one
- ◯ Yes, more than one

57. What is the exact name of your neurological condition? *

*If you suffer from several conditions, please consider only the one which is the most severe*

58. What is the classification of your neurological condition? *

*If you suffer from several conditions, please consider only the one which is the most severe*

- ◯ Movement disorders (Parkinson’s disease, Tic disorders, Ataxic disorders)
- ◯ Disorders with neurocognitive impairment as a major feature (Alzheimer’s disease, Lewy body disease)
- ◯ Multiple sclerosis or other white matter disorders (Neuromyelitis optica, Leukodystrophies)
- ◯ Epilepsy or seizures
- ◯ Headache disorders (Migraine)
- ◯ Cerebrovascular diseases (Stroke, Cerebral ischaemia)
- ◯ Spinal cord disorders excluding trauma (Myelopathy, Myelitis, Cauda equina syndrome)
- ◯ Motor neuron diseases or related disorders (Spinal muscular atrophy)
- ◯ Disorders of nerve root, plexus or peripheral nerves (Radiculopathy)

- ◯ Diseases of neuromuscular junction or muscle (Myasthenia gravis)
- ◯ Cerebral palsy
- ◯ Nutritional or toxic disorders of the nervous system (caused by obesity or alcohol-use)
- ◯ Disorders of cerebrospinal fluid pressure or flow (Hydrocephalus)
- ◯ Disorders of autonomic nervous system
- ◯ Human prion diseases (Creutzfeldt-Jacob disease)
- ◯ Disorders of consciousness
- ◯ Postprocedural disorders of the nervous system (caused by surgery or medical process)
- ◯ Injuries of the nervous system
- ◯ Neoplasms of the nervous system
- ◯ Structural developmental anomalies of the nervous system
- ◯ Syndromes with central nervous system anomalies as a major feature
- ◯ Paralytic symptoms (Tetraplegia, Hemiplegia)
- ◯ Dissociative neurological symptom disorder
- ◯ I do not know
- ◯ Other

59. If you answered "Other" to the previous question, could you specify?

60. For how long have you suffered from this neurological condition (from the diagnosis, in years)? *

*If you suffer from several conditions, please consider only the one which is the most severe*

The value must be a number

61. Do you receive treatment for this neurological condition? *

*If you suffer from several conditions, please consider only the one which is the most severe*

- ◯ No, none
- ◯ Yes, drugs only
- ◯ Yes, surgery only
- ◯ Yes, brain stimulation (other than drugs)
- ◯ Yes, both drugs and surgery
- ◯ Yes, both drugs and brain stimulation (other than drugs)
- ◯ Yes, drugs, surgery and brain stimulation (other than drugs)
- ◯ Other

62. If you answered "Other" to the previous question, could you specify?

### Thank you!

Thank you very much for your time and your answers.
Your answers will help us to improve our understanding about how neuromodulation is perceived.
Your data will be exclusively used for a scientific research objective.

Please, feel free to share this survey as much as possible.

You can share this link with all your contacts:
**---> https://forms.office.com/r/YNhmVAYYFA <---**

If you are happy to be updated about our research or be contacted for further studies, please enter your details in the following separate form: https://forms.office.com/r/Tb4bcTpT3X

If you have any query, feel free to contact the following research associate:


**Please do not forget to click on the "send" button bellow to validate your answers.**

This content is neither created nor endorsed by Microsoft. The data you submit will be sent to the form owner.

Microsoft Forms
